# The utility of Resting Pulse Rate in Defining High Blood Pressure among Adolescents in Mbarara Municipality, Uganda

**DOI:** 10.1101/2020.07.14.20153387

**Authors:** Godfrey Katamba, David Collins Agaba, Rosemary Namayanja, Agnes Namaganda, Abdul Musasizi, Mivule Abdul Kinene, Richard Migisha

**Affiliations:** Department of Physiology, School of Medicine, King Ceasor University, Kampala, Uganda; Department of Physiology, Faculty of Medicine, Mbarara University of Science and Technology, Mbarara Uganda; Department of Biochemistry, School of Medicine, King Ceasor University, Kampala, Uganda; Department of Human Anatomy, School of Medicine, King Ceasor University, Kampala, Uganda

**Author notes:** Corresponding author; Godfrey Katamba, King Ceasor University, Kampala Uganda; email.

## Abstract

High resting pulse rate (RPR) is associated with adverse cardiovascular events, and could be used as a marker of cardiovascular health. We determined the correlation between RPR and blood pressure (BP); and its accuracy in defining high blood pressure among adolescents attending secondary schools in Mbarara municipality, south-western Uganda. We conducted a cross-sectional study among secondary school adolescents aged 12-19 years in Mbarara municipality, Uganda. We captured demographic characteristics using a structured questionnaire; and measured anthropometric indices and BP. We performed a linear regression analysis to determine the relationship between RPR and blood pressure and plotted receiver operating characteristics curves (ROC) to assess the accuracy of RPR in defining high BP. We enrolled 616 adolescents with a mean age of 15.6±2.0 years and 65.6% (404/616) were female. The RPR was significantly correlated with diastolic blood pressure (DBP) in both boys (Beta = 0.22 [95%CI: 0.10; 0.36]), p<0.001 and girls (Beta = 0.51 [95%CI: 0.43; 0.60]), p<0.001. RPR was significantly correlated with systolic blood pressure (SBP) only in the girls (Beta = 0.23 [95%CI: 0.15; 0.30]), p<0.001. The optimal threshold for RPR in defining prehypertension was RPR≥76bpm with an area under the curve (AUC) of 0.653[95%CI: 0.583-0.722], the sensitivity of 0.737 and specificity of 0.577. In defining hypertension, the optimal threshold was RPR ≥ 79bpm at a sensitivity of 0.737 and specificity of 0.719, with an AUC of 0.728[95%CI: 0.624-0.831]. Resting pulse rate was positively correlated with BP and was more accurate in defining hypertension compared to prehypertension in the study.

## Introduction

Hypertension (HTN) was previously considered a rare condition among adolescents^1^. In contrast, it has become more common and is increasing unexpectedly in this age group ^2,3^. The prevalence of HTN among adolescents has been reported in several countries and varies from 3.6% to 30% ^3-11^. High blood pressure (BP) in young subjects is associated with accelerated vascular aging and increased risk for adult HTN and cardiovascular disease^12,13^. The diagnostic algorithm for HTN in children and adolescents does not rely on single BP cut-offs as is the case for adults. This is believed to be the cause of its under-diagnosis in young populations, because the normal and abnormal BP values vary with sex, height, and age, making it difficult for clinicians to remember^14^. Furthermore, the algorithm involves repeated BP measurements and follow up visits^15,16^. This is laborious and may not be suitable for resource-limited settings, with a low clinician to patient ratio. Thus, easier to use diagnostic approaches are needed in resource-limited settings, to facilitate the identification of adolescents with high BP and those who need interventions to prevent or control HTN to improve long-term outcomes.

Resting pulse rate (RPR) has recently attracted attention as an important predictor of high BP^17,18^ and a risk factor for cardiovascular morbidity and mortality in general populations^19-22^. Additionally, various studies among children and adolescents, have reported a significant correlation of elevated RPR with both systolic (SBP) and diastolic blood pressure (DBP)^23-26^, highlighting its possible role in screening for high BP. Hypertension results from alterations in the overall hemodynamic load on the cardiovascular system^27^, which is an interplay between the cardiac output (CO) and total peripheral vascular resistance (TPR)^28^. The CO is affected by changes in the heart rate, which in turn depends on the autonomic tone of the cardiovascular system^29^. The relationship between RPR and BP, among adolescents, has not been well studied in sub-Saharan Africa. RPR is considered a simple and cheap measurement, which does not necessitate the use of special instruments and patients can conveniently measure it by themselves.

In this cross-sectional study, we aimed to assess the utility of RPR in defining high blood pressure (SBP and DBP) among adolescents from selected secondary schools in Mbarara municipality. South-western Uganda.

## Methods

### Study sample

The participants were part of a cross-sectional study ((IRB No. 18/03-18), which involved three secondary schools in Mbarara municipality, southwestern Uganda between May and November 2018. The sample size was estimated using the Kish Leslie method of 1965 ^30^ assuming a prevalence of hypertension of 10.7% among secondary school adolescents ^31^, and 95% confidence interval within a 3% error margin. The final sample size was adjusted for an anticipated participant non-response rate of 10%; thus 449 participants were required. In this analysis, a total sample of 616 adolescents aged 12-19 years was used.

### Socio-demographic information

The age of the participant was self-reported and was the number of complete years since birth while the sex of the participant was a binary variable (male or female) and based on the sexual characteristics as was observed by the researcher.

### Determination of anthropometric indices

The methods used to determine the height, weight, neck circumference, waist circumference, and their respective ratios, have been described in detail elsewhere ^32^. Briefly, height was measured using a wall mount height board in centimeters with the participant having no shoes ^6^. A standard Seca scale was used to determine the weight to the nearest 0.5kg ^33^. Body mass index (BMI) was calculated as the ratio of weight in kilograms to height in square meters. Waist circumference (WC) was measured at the midpoint between the lowest border of the rib cage and the top of the lateral border of the iliac crest while hip circumference was measured at the greatest horizontal circumference below the iliac crest at the level of the greater trochanter using a non-elastic measuring tape (Seca 203 Ergonomic circumference measuring tape, Hamburg, Germany). Waist to hip ratio (WHR) was the ratio of WC to HC ^34^ while waist to height ratio (WHtR) was the ratio of WC divided by height ^35^. The Neck circumference (NC) assessed as a surrogate measure for upper body adipose tissue distribution. It was measured at the level of the laryngeal prominence using an inelastic flexible measuring tape (Seca 203 Ergonomic circumference measuring tape, Hamburg, Germany), with the subjects in the standing position and the head held erect and eyes facing forward to the nearest 0.1cm ^36^.

### Measurement of blood pressure and resting pulse rate

Blood pressure and resting pulse rate were measured using a digital blood pressure machine (Scian SP-582 Digital Blood Pressure Monitor) as previously described by Katamba et al ^32^. Each participant was allowed to seat on a chair with back supported, feet on the floor, arm supported and cubital fossa at heart level after 5 minutes of sitting rest without talking ^37,38^. The cuff of appropriate size (ranged from 12×22cm to16×30cm) was placed at the bare upper arm, one inch above the bend of the participant’s elbow. It was ensured that the tubing fell over the front center of the arm so that the sensor was correctly placed. The end of the cuff was pulled so that it was evenly tight around the arm. The cuff was placed tight enough so that only two fingertips could be slipped under the top edge of the cuff. It was made sure that the skin did not pinch when the cuff inflated. The participant was asked to remain and quiet as the machine begins measuring. The cuff inflated automatically after placing the start button, and then slowly deflated so that the machine took the measurement. When the reading was complete, the monitor displayed the BP and pulse rate on the digital panel ^39^. Three readings were recorded per participant at 5 minutes interval. The average of the 2nd and 3rd SBP and DBP measurements was used as the subject’s BP respectively ^38^. Those adolescents who had elevated BP in the first session were identified. A re-measurement, using the same procedure was done after one week to confirm that BP is truly and constantly elevated.

### Data management and analysis

Data were analyzed using the Stata software version 13.0 (College Station, Texas, USA). Continuous variables such were described as mean±SD while categorical variables such as sex were described as percentages and frequencies. The outcome variable was BP (both SBP and DBP). Pearson correlation analysis was done to determine the strength of the relationship between BP, RPR, and anthropometric indices. The Pearson correlation coefficients and their 95% confidence intervals were reported. Multivariate linear regression analysis was used to control for anthropometric indices in the relationship between RPR and BPs. A p-value of less than 0.05 was considered for assessing the statistical significance and 95% CI of the changes in BP values associated with a unit change in RPR was calculated. Receiver operating characteristic (ROC) curve analyses were performed to determine the accuracy of RPR in defining high BP (prehypertension and hypertension) among adolescents and identify optimal thresholds of RPR for identifying high arterial BP. Optimal thresholds were selected as the values corresponding to the maximum of Youden’s index on the ROC curve. Prehypertension and hypertension were then redefined by the determined optimal thresholds of RPR. These were compared with the gold standard blood pressure cut-offs in adolescents as stated by the Joint National Committee on hypertension guidelines ^40^. The sensitivities and specificities and their respective 95% confidence intervals (CI) were obtained to assess the performance of the determined RPR optimal thresholds. All analyses were stratified by sex because of the physiological difference in resting pulse rates between the two groups.

## Results

### Characteristics of study participants by body mass index

A sample of 616 had complete data and was included in the final analysis as was obtained from a larger prevalence study. These were aged between 12 to 19 years; with a mean age of 15.6±2.0 years. The prevalence of hypertension was found at 3.1% while prehypertension was estimated at 7.1% ^41^. In table 1, the characteristics of the participants are described according to BMI categories.

**Table 1:** Showing characteristics of participants by BMI categories

| Variable | Obese<br>(n=21) | Overweight<br>(n=188) | Normal weight<br>(n=387) | Underweight<br>(n=20) |
| --- | --- | --- | --- | --- |
| **Female** | (n=18) | (n=166) | (n=215) | (n=5) |
| **Male** | (n=3) | (n=22) | (n=172) | (n=15) |
|  | *Mean (SD)* | *Mean (SD)* | *Mean (SD)* | *Mean (SD)* |
| Age (years) | 17.7(1.6) | 15.6(1.9) | 15.5(2.0) | 15.9(1.9) |
| NC (cm) | 31.9(1.8) | 30.3(1.7) | 29.9(2.2) | 28.8(1.6) |
| WHR | 0.84(0.05) | 0.76(0.07) | 0.8(0.08) | 0.79(0.07) |
| RPR (bpm) | 77.9(6.8) | 74.9(7.0) | 75.6(8.7) | 76.4(13.5) |
| SBP (mmHg) | 117.3(14.6) | 114.3(7.3) | 112.9(8.9) | 107(14.4) |
| DBP (mmHg) | 75.3(7.9) | 65.8(7.4) | 66.4(8.1) | 66.4(9.7) |

SBP=Systolic blood pressure, DBP=Diastolic blood pressure, NC=neck circumference, WHR=waist height ratio, RPR=resting pulse rate

### Correlation analysis

RPR was positively correlated with both SBP and DBP among adolescents in our study. While the overall linear correlation coefficients of 0.22 [95% CI: 0.14 – 0.29] for SBP and 0.44 [95% CI: 0.37 – 0.50] for DBP were statistically significant, they are weak. The correlation of RPR with both SBP and DBP was still significant and positive even after controlling for the sex of participants as shown in figure 1. Most of the indices showed positive linear relations with both DBP and SBP among the girls, except for the WHR, which exhibited a negative correlation with SBP as seen in table 2.

**Table 2:** Showing the correlation between blood pressure and anthropometric indices

| Variables | r (Pearson correlation)<br>[95% confidence interval] | p-value |
| --- | --- | --- |
| **Systolic blood pressure** |  |  |
| BMI (kgm<sup>-2</sup>) | 0.10 [0.02 - 0.18] | 0.011 |
| NC (cm) | 0.50 [0.44 - 0.56] | <0.001 |
| WHR | -0.14[-0.21-0.39] | <0.001 |
| WHtR | 0.24 [0.17 - 0.31] | <0.001 |
| **Diastolic blood pressure** |  |  |
| BMI (kgm<sup>-2</sup>) | 0.05 [0.03 - 0.12] | 0.265 |
| NC (cm) | 0.32 [0.25 - 0.39] | <0.001 |
| WHR | -0.14[-0.22-0.07] | <0.001 |
| WHtR | 0.15 [0.07 - 0.23] | <0.001 |

BMI=Body mass index, NC= Neck circumference, WHR= Waist hip ratio, WHtR=Waist height ratio.

**Figure 1:**
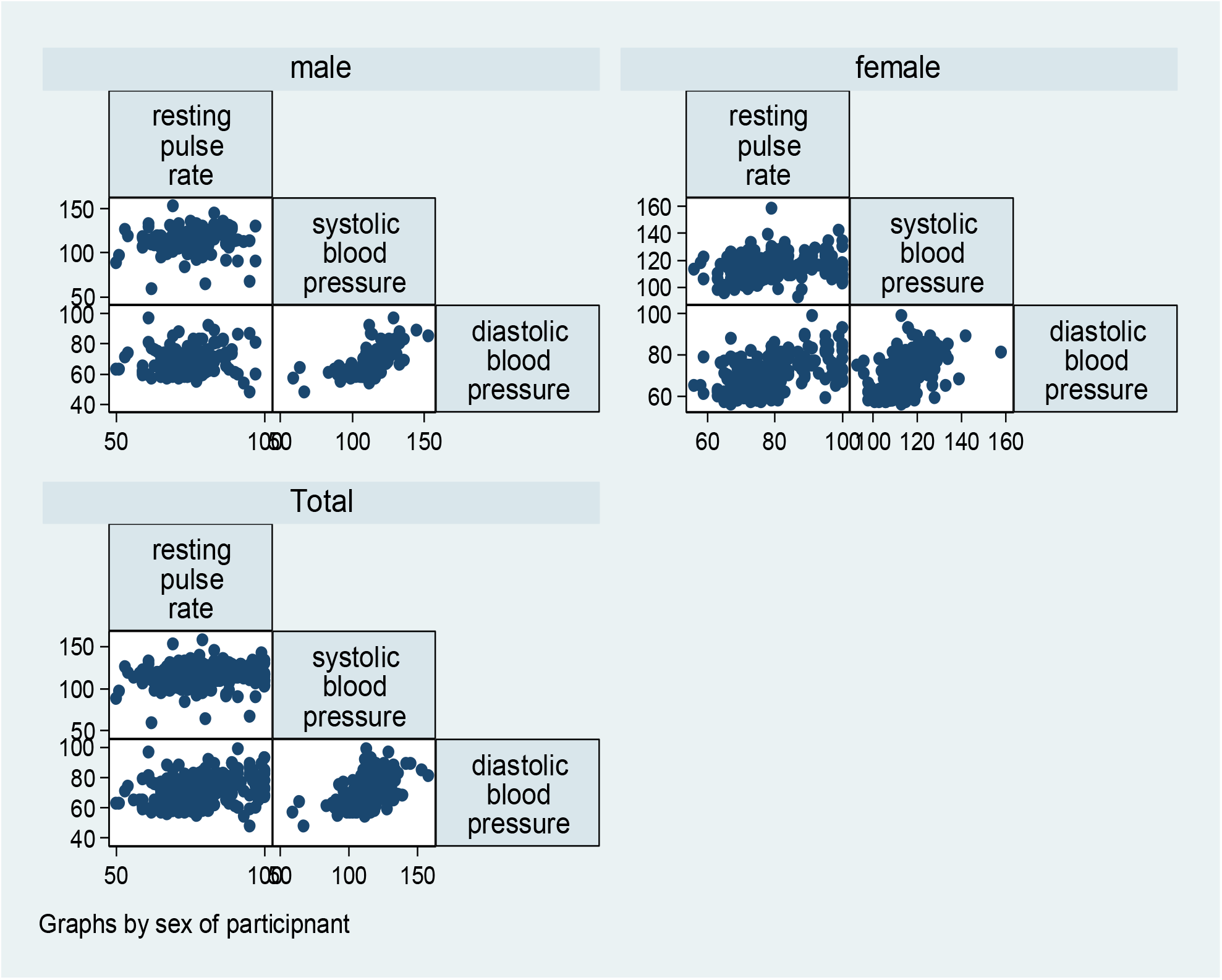
Showing the Pearson correlation of resting pulse rate with blood pressure of the participants

### Linear regression analysis

A bivariate linear regression of RPR and BP (SBP and DBP) indicated a significant positive linear relationship between two variables. However, the variability (using the linear regression R squared value) in DBP, which could be explained by RPR, was only 19% compared to 5% for SBP. For the entire sample, a multivariate linear regression showed that a unit increase in RPR was significantly associated with increases in BP (SBP and DBP). The change in SBP reduced after adjusting for anthropometric indices as shown in table figures 2a and 2b and figures 3a and 3b.

**Figure 2a:**
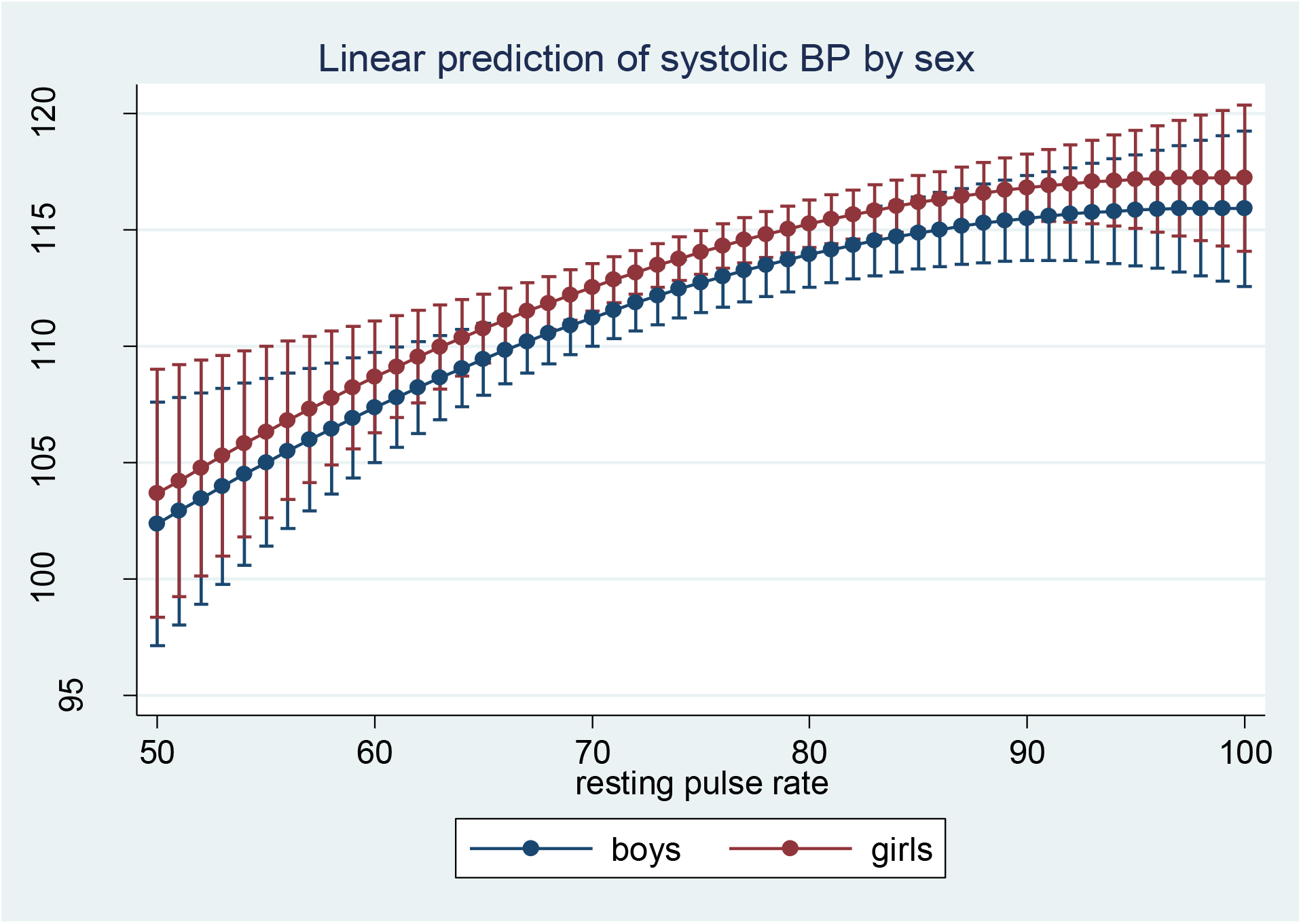
Showing linear regression of resting pulse rate with systolic blood pressure by sex of the participants

**Figure 2b:**
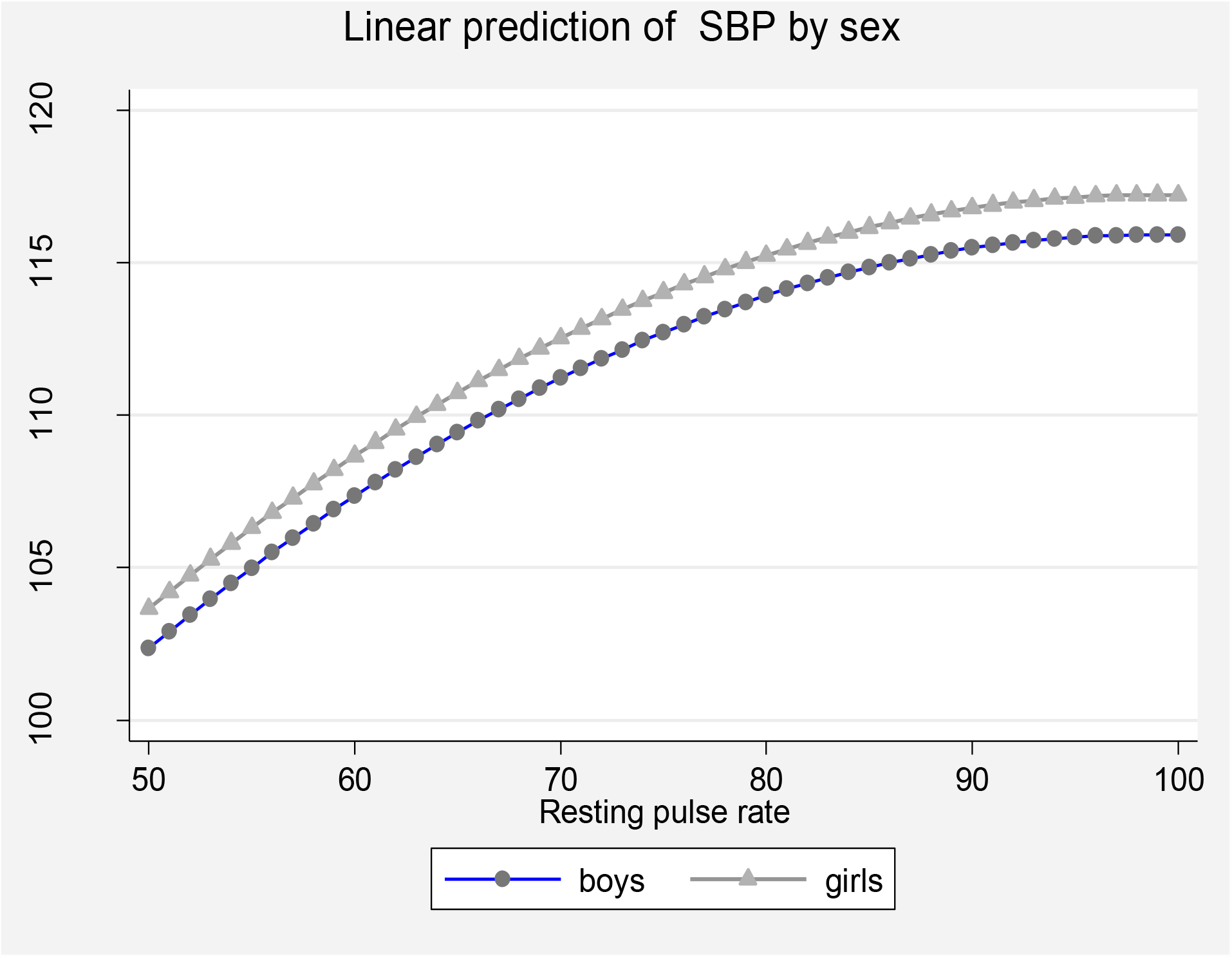
Adjusted linear regression of resting pulse rate with systolic blood pressure by sex of the participants

**Figure 3a:**
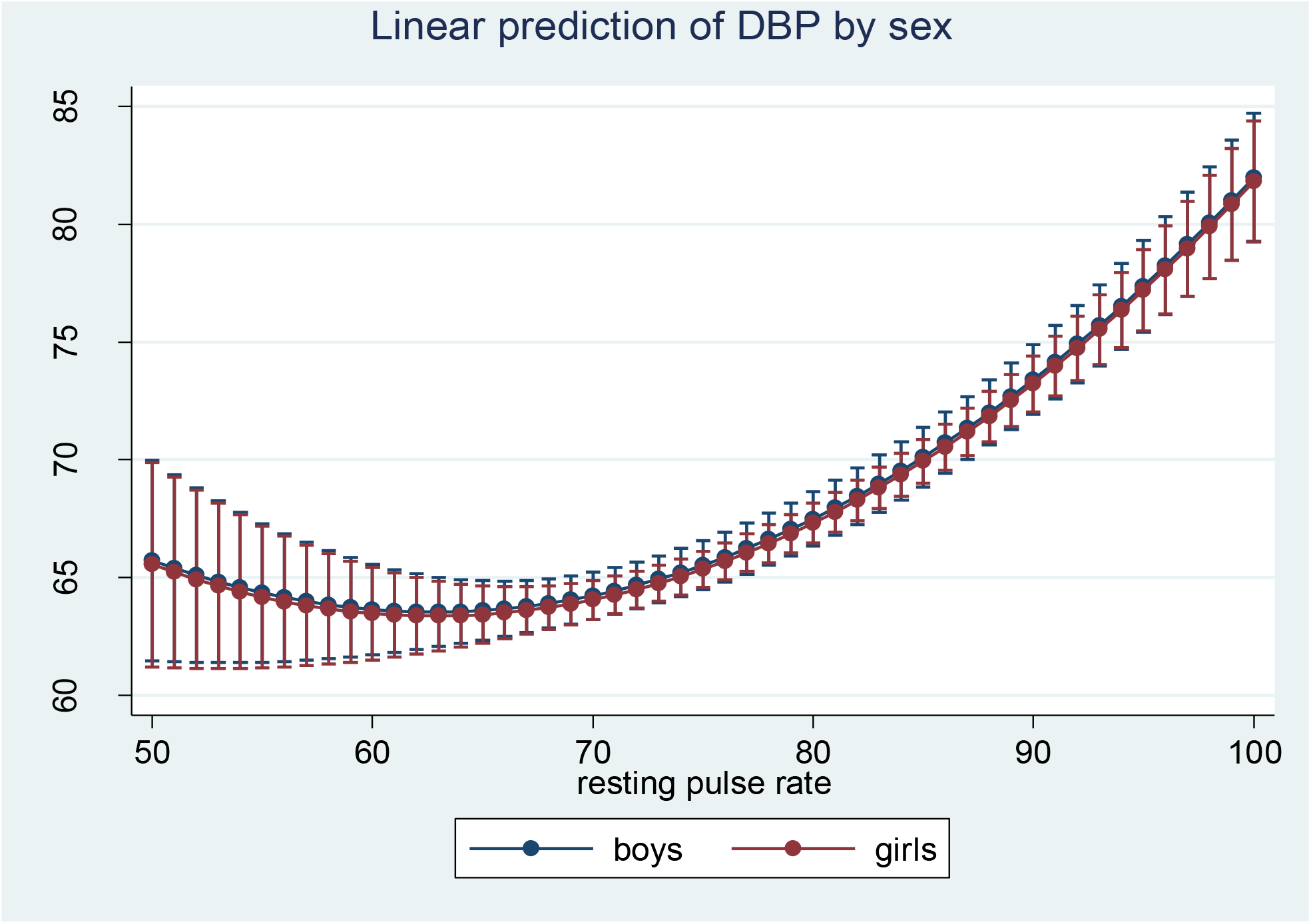
Showing linear regression of resting pulse rate with systolic blood pressure by sex of the participants

**Figure 3b:**
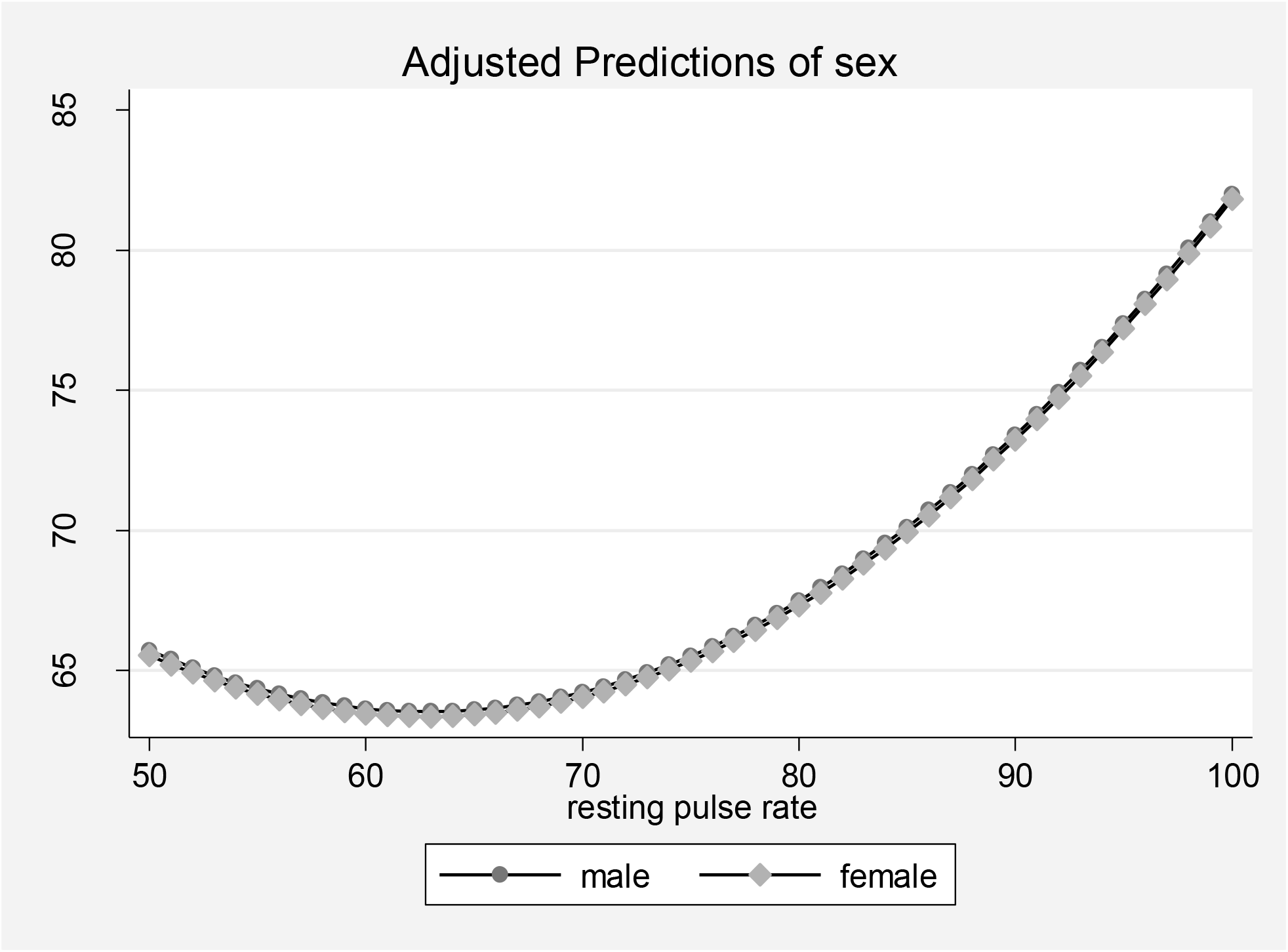
Adjusted linear regression of resting pulse rate with diastolic blood pressure by sex of the participant.

### Optimal thresholds of RPR for identifying prehypertension and hypertension among adolescents

The optimal threshold for identifying prehypertension was determined as RPR≥76bpm with a sensitivity of 0.737 and specificity of 0.577. The performance of this threshold based on the AUC was 0.652 [CI; 0.583-0.722] as shown in figure 4. This optimal threshold classified 58.8% of the participants correctly as prehypertensive. The positive predictive value (PPV) of 0.118 and a negative predictive value (NPV) of 0.882. The optimal threshold for hypertension in our study was determined as RPR ≥79 bpm, with a sensitivity of 0.737, the specificity of 0.719 and an AUC of 0.728[CI; 0.624-0.831] as shown in figure 5. This threshold classified 71.92% of the participants correctly, with a PPV of 0.077 and NPV of 0.988. Both sensitivity and specificity of RPR were higher for the identification of HTN than prehypertension among our study participants as shown in table 3.

**Table 3:** Optimal thresholds of RPR for identifying PreHTN and HTN among adolescents

| Category | Threshold | PPV | NPV | Se | Sp | AUC [95% CI] |
| --- | --- | --- | --- | --- | --- | --- |
| **Overall** |  |  |  |  |  |  |
| PreHTN | ≥76bpm | 0.117 | 0.965 | 0.727 | 0.577 | 0.653[0.583-0.722] |
| HTN | ≥79bpm | 0.077 | 0.989 | 0.737 | 0.719 | 0.728[0.624-0.831] |
| **Boys** |  |  |  |  |  |  |
| PreHTN | ≥75bpm | 0.167 | 0.975 | 0.722 |  | 0.685[0.558-0.811] |
| HTN | ≥76bpm | 0.154 | 0.993 | 0.700 | 0.608<br>0.629 | 0.674[0.470-0.877] |
| **Girls** |  |  |  |  |  |  |
| PreHTN | ≥76bpm | 0.141 | 0.966 |  |  | 0.623[0.515-0.731] |
| HTN | ≥78bpm | 0.053 | 0.100 | 0.769<br>0.100 | 0.545<br>0.620 | 0.822[0.739-0.905] |

**PreHTN:** Prehypertension, **HTN:** Hypertension, **Se:** sensitivity, **Sp:** Specificity, **PPV:** positive predictive value, **NPV:** Negative predictive value, **AUC:** area under the curve.

**Figure 4:**
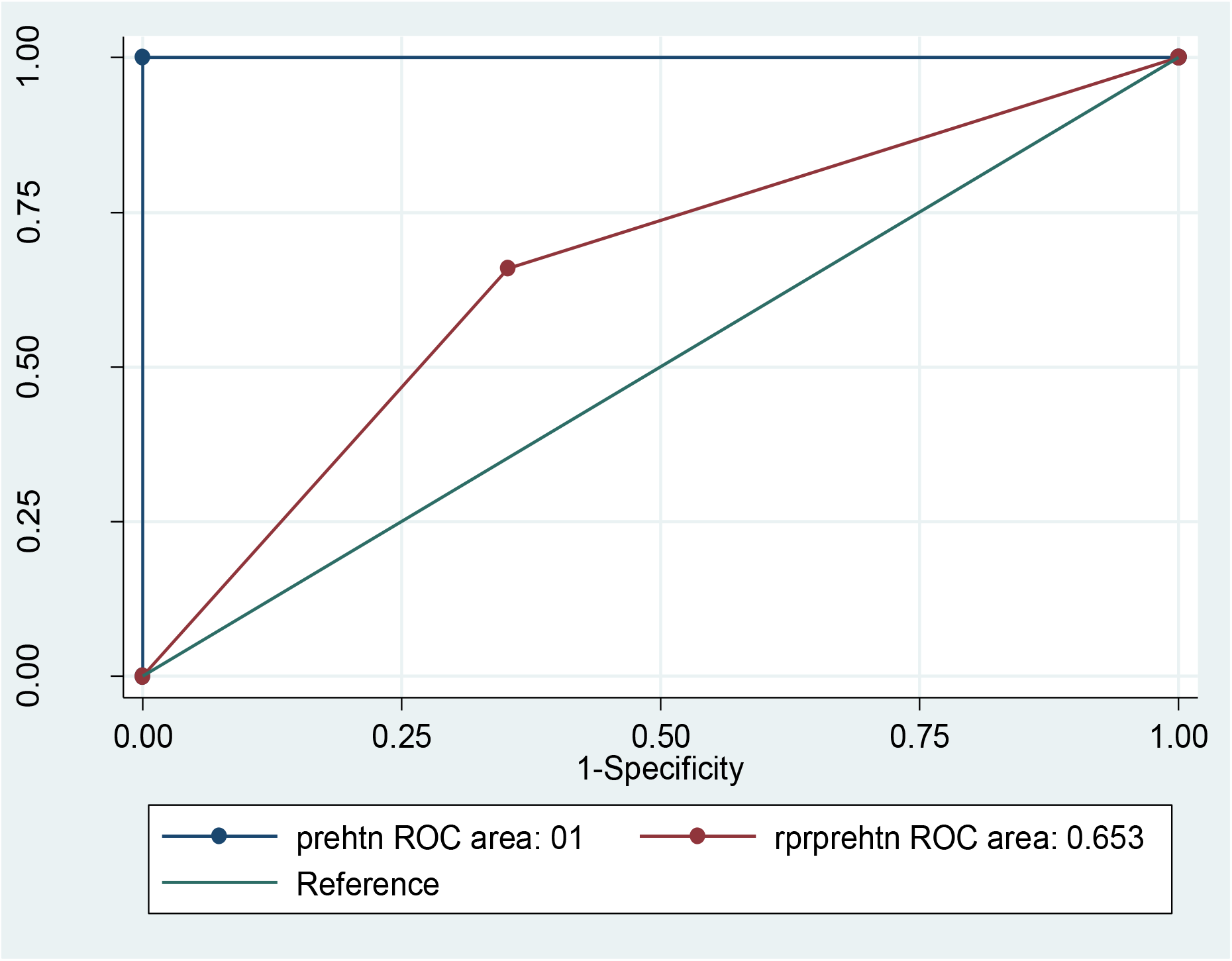
ROC curve sowing the overall prehypertension definition by resting pulse rate among adolescents. prehtn: *prehypertension*, rprprehtn: resting *pulse rate in defining prehypertension*

**Figure 5:**
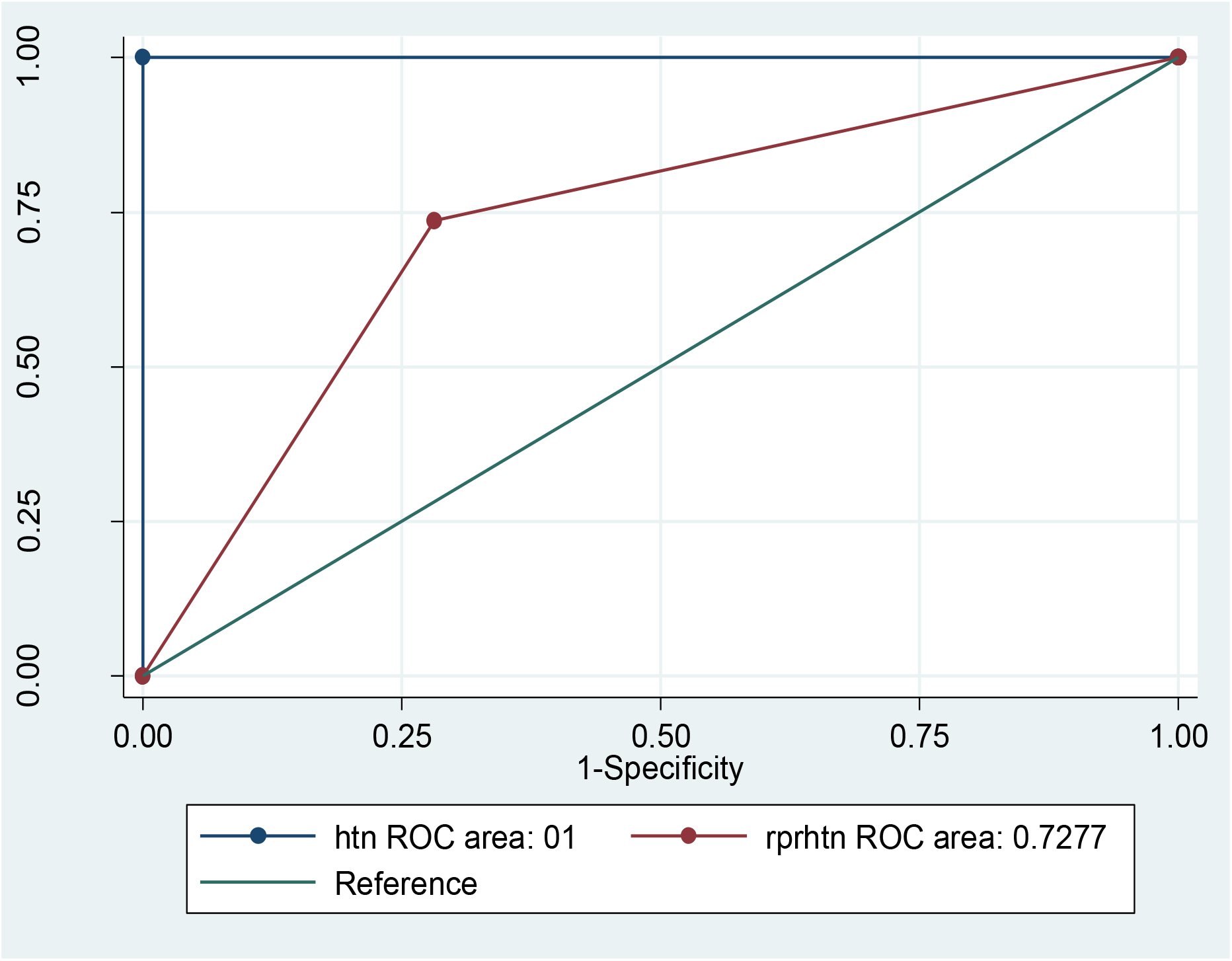
ROC curve showing the overall AUC of resting pulse rate in defining hypertension among adolescents. htn: *hypertension*, rprhtn: *resting pulse rate in defining hypertension*

## Discussion

Our study demonstrated a positive linear relationship between RPR and blood pressure (SBP and DBP), even after adjusting for anthropometric indices and sex. This is in agreement with findings from several studies done among children and adolescents in Brazil^42,43^, Nigeria ^31^, India ^44^, USA ^45^, and China ^25,46,47^. The association between RPR and elevated BP is believed to be due to alterations in the cardiovascular autonomic tone, including a reduction in cardiac vagal stimulation, increased sympathetic activation and imbalance in the sympathovagal system^48^. As previously reported^49,50^, these alterations are associated with the recognized increased risk for HTN among individuals with elevated resting heart rates. Moreover, it has been previously postulated that the effect of elevated heart rate on cardiovascular mortality might be mediated through high BP^26^.

Our study also found RPR to be more accurate in defining HTN as opposed to defining prehypertension among adolescents. The AUC of 0.609 to 0.854 suggested a high and robust discriminatory accuracy of RPR in identifying HTN among secondary school adolescents in our study setting. In defining prehypertension, the AUC was lower but well above 0.6 which is the recommended minimum. Besides, the much higher NPVs of the cut off for HTN indicated that our test is unlikely to omit adolescents with HTN and prehypertension. However, due to the low prevalence of HTN in our study, the PPVs of our optimal cut-offs were also low. This showed that many adolescents with both HTN and PreHTN would be misclassified as not having prehypertension or hypertension. This makes RPR not the best substitute for the commonly used age, gender and height BP percentiles for diagnosing hypertension. However, it can be used for screening adolescents at high risk of high BP in populations with a high prevalence of high BP and in resource-limited settings. The study findings are in agreement with those as reported by Tjugen *et al* ^51^, which involved elderly subjects to determine whether the heart rate itself was a risk factor for the development of hypertension or just a marker of sympathetic overactivation. It was found that a high heart rate was a strong predictor of cardiovascular disease and a reliable predictor for the development of hypertension ^51^. The study also addresses recommendations for further research on the use of RPR in defining HTN among adolescents from China ^46^, Hong Kong ^25^ among others. Thus, this study responds to such recommendations with fair news, showing that RPR may be useful in discriminating people who are at risk of developing elevated BP; however, it may not be used as the basis for the diagnosis of the condition. Nonetheless, our findings contribute to the existing body of evidence that proposes the use of RPR as a useful clinical measurement and as a risk factor for cardiovascular disease as was reported from the WHO Cardiovascular Disease and Alimentary Comparison Study ^23^.

Our study has some limitations as outlined; First, the participants were not objectively evaluated secondary causes of high blood pressure and elevated heart rate. Nevertheless, we did not exclude adolescents with a self-reported history of other endocrine disorders and those with acute febrile conditions. Additionally, we assessed adolescents with high BP and elevated RPR on more than one occasion before confirming the diagnosis. Second, we conducted the study in a few selected secondary schools, because of logistical concerns. This limits the generalization of our findings to adolescent populations outside the peri-urban secondary schools of Mbarara municipality, south-western Uganda. We thus recommend larger surveys in a wider population to corroborate our findings. Finally, we cannot assess the temporal relationship between RPR and BP in our study population, because of the cross-sectional nature of our study design. Future longitudinal studies in larger adolescent populations are needed to explore this relationship.

## Conclusions

Our study demonstrated a positive linear relationship between RPR and BP among adolescents after controlling for anthropometric indices and sex. RPR was found to be more accurate in defining hypertension than in defining prehypertension in our study population.

## Data Availability

Data is available on request from the corresponding author

## List of abbreviations

AUC: area under the curve
BMI: Body mass index
BP: blood pressure
CI: Confidence interval
CO: cardiac output
CVD: Cardiovascular disease
DBP: diastolic blood pressure
HTN: Hypertension
NC: Neck circumference
NPV: negative predictive value
PPV: positive predictive value
PreHTN: Prehypertension
ROC: receiver operating characteristic
RPR: resting pulse rate
SBP: systolic blood pressure
TPR: total peripheral resistance
WHR: Waist hip ratio
WHtR: Waist height ratio

## Declarations

### Ethical approval and consent to participate

The study was approved by the research ethics committee of Mbarara University of Science and Technology (IRB No. 18/03-18) We also obtained permission to collect data from the school headteachers. The class teachers were informed about the purpose of the study and all potential participants were first sensitized about study procedures, possible benefits, and risks. Adolescents of 12 – 17 years freely assented, and consent for their participation was obtained through their teachers. We obtained written informed consent from adolescents aged 18-19 years.

### Consent for publication

Not applicable

### Availability of data and materials

The dataset is available on request from the corresponding other

### Authors’ contributions

GK, RM, and DCA: Conceptualization of work & its realization, wrote the manuscript, checked the references, compiled the literature sources, data collection, statistical analysis, and interpretation of data, and wrote the manuscript and is the corresponding author.

RN, DCA, RM: mentored the conceptualization of work & its realization, compiling literature sources and statistical analysis, helped in data interpretation, guided manuscript writing, checked the references.

AN, AM, MAK: Assent and consent form administration, data collection, data entry, and analysis. All authors read and approved the study manuscript

## Acknowledgments

I am thankful to my family, friends and the participants from the various data collection sites.

